# Socioeconomic determinants and spatial distribution of leprosy in São Paulo State, Brazil (2017–2020): a municipal-level analysis

**DOI:** 10.1101/2025.09.16.25335912

**Authors:** Carlos Souto dos Santos Filho, Ana Júlia Alves Câmara, Guilherme Aparecido Santos Aguilar

## Abstract

**Objectives:** Leprosy remains a significant public health challenge in Brazil, with distinct spatial and temporal patterns reflecting underlying socioeconomic inequalities. This study investigated the association between municipal socio-territorial indicators and the geographic distribution of leprosy in São Paulo State.

**Methods:** A total of 4562 new leprosy cases reported across 645 municipalities from 2017 to 2020 were analysed using Bayesian spatial modelling. The Besag–York–Mollié 2 (BYM2) model was implemented to estimate relative risk while accounting for spatial dependence, incorporating municipal wealth as a covariate. Expected case counts were calculated based on overall state incidence rates and local population sizes. Exceedance probabilities were computed to identify high-risk clusters using a threshold of relative risk >2.0.

**Results:** Local relative risk estimates ranged from 0.06 to 31.52 (median: 0.96), with substantial spatial heterogeneity across municipalities. Municipal wealth demonstrated a significant protective effect (posterior mean: −0.059, 95% CrI: −0.081 to −0.037), indicating lower leprosy incidence in economically advantaged areas. The mixing parameter (*φ* = 0.517) revealed approximately equal contributions of structured spatial and unstructured random effects. High-risk clusters were predominantly concentrated in western and northern regions of the state.

**Conclusions:** Leprosy distribution in São Paulo State exhibits significant spatial clustering associated with municipal wealth disparities. The persistent geographic patterns suggest that socioeconomic conditions are associated with variations in local leprosy risk across municipalities. These findings support targeted public health interventions in identified high-risk areas and highlight the importance of addressing underlying sociodemographic inequalities that may influence the local effectiveness of leprosy control strategies.

## Introduction

Hansen’s disease, caused by *Mycobacterium leprae* and *Mycobacterium lepromatosis*, is a chronic infectious disease ranking as the second most common mycobacterial infection world-wide after tuberculosis.^1^ The bacteria target Schwann cells and dermal macrophages, causing peripheral neuropathy, cutaneous manifestations, and mucosal lesions that may progress to motor and sensory impairments, deformities, and stigmatising wounds. In addition to its clinical impact, historically, the disease carries profound social implications due to historical discrimination and exclusion.

Key scientific milestones include Hansen’s discovery of *M. leprae* in 1873; Shepard’s successful bacterial cultivation in mouse footpads at the U.S. Centers for Disease Control in 1959; and Kirchheimer and Storrs’ development of the nine-banded armadillo model in 1968, which enabled disseminated infection studies. These advances significantly enhanced understanding of the disease’s pathogenesis and treatment approaches.^2^

Despite decades of public health initiatives, leprosy continues to affect populations disproportionately in low- and middle-income countries (LMICs), where poverty, poor housing conditions, and limited access to health care persist. Globally, from 1990 to 2021, the age-standardised incidence rate (ASIR) for leprosy declined by 64.79%, reaching 0.58 per 100,000 population in 2021.^3^ The global burden, as measured by disability-adjusted life years (DALYs), also followed a downward trajectory, yet leprosy remains endemic in more than 120 countries. Notably, Brazil, India, and Indonesia together account for approximately 79.3% of new annual cases, with the disease particularly concentrated in tropical and subtropical regions.^1^ In these contexts, the persistence of leprosy reflects not only biological transmission mechanisms but also structural and societal inequities that facilitate the disease*’*s spread and hinder timely diagnosis and treatment.

In Brazil, leprosy continues to represent a complex public health issue, with distinct spatial and temporal patterns. Recent investigations have documented heterogeneous relapse rates and disease clustering across the national territory. Using space–time scan statistics, Boigny and colleagues identified high-risk clusters of leprosy relapse in the North, Northeast, and Central-West regions, characterised by relative risks (RR) ranging from 1.78 to 3.97.^4^ These clusters were frequently found in socioeconomically disadvantaged municipalities with barriers to health service access. According to the authors, these patterns are associated with both structural vulnerabilities and higher baseline infection rates, which contribute to continued transmission and recurrence of the disease.

In a complementary analysis, Sczmanski *et al*.^5^ assessed clinical and epidemiological data from 215,155 newly reported leprosy cases between 2014 and 2019. Their findings revealed that the disease remains stable in most Brazilian regions, with the highest detection rates in the Midwest and lowest in the South. The most affected individuals were predominantly male, literate, and of black or brown ethnicity, aged between 60 and 79 years. Multibacillary forms and borderline clinical presentations predominated, suggesting late detection and heightened transmission potential. The study emphasised that leprosy persists as a public health concern, closely linked to social inequalities and requiring spatially informed public health responses. For a long time, the importance of epidemiology related to leprosy was neglected in spatial studies. Guided by these findings, the present study adopts a spatial epidemiology approach to investigate the association between municipal-level socio-territorial indicators and the geographic distribution of leprosy in the state of São Paulo from 2017 to 2020. By incorporating spatial statistical modelling and disease mapping techniques, this analysis aims to uncover hidden patterns of risk and support the strategic allocation of public health resources in line with the WHO’s “Towards Zero Leprosy” roadmap.

The main objective of this study was to evaluate the association between municipality-level socio-territorial indicators—namely wealth, longevity, and education—and the spatial distribution of leprosy cases in the State of São Paulo from 2017 to 2020. Additionally, the study aimed to identify clusters of municipalities where the incidence of leprosy was significantly higher than expected based on available covariates, suggesting the presence of underlying social or environmental determinants not captured by standard indicators. Furthermore, the study sought to compare patterns of leprosy incidence across pre-defined socio-territorial groups of municipalities, with a particular focus on assessing whether combinations of low wealth were consistently associated with increased leprosy risk.

## Materials and methods

### STUDY AREA: STATE OF SÃO PAULO

Demographic data from the 2022 Brazilian Census indicate that the state of São Paulo reached a total population of 44.4 million, reflecting an increase of 3.2 million residents over the previous twelve years, corresponding to an average annual growth of 263,200 individuals. Since 1940, the population has expanded by a factor of 6.2.^6^ However, recent decades have witnessed a deceleration in growth rates due to declining fertility and a reduction in net migration.

Between 2010 and 2022, the capital’s annual growth rate was 0.15%, significantly lower than the rates observed in the interior (0.79%) and the state as a whole (0.62%). The most recent census confirmed a reduction in the state’s overall population growth rate, which fell from 1.09% in the 2000–2010 period to 0.62% in the subsequent intercensal period. This shift was accompanied by a rise in the number of municipalities experiencing population decline, increasing from 102 to 197, while the number of municipalities with positive growth declined from 543 to 448.^6^

In addition to demographic indicators, the São Paulo Index of Social Responsibility (IPRS), which evaluates municipalities based on wealth, longevity, and educational attainment, revealed notable changes between 2014 and 2018. The number of municipalities classified as vulnerable, characterised by low levels in all three dimensions, decreased from 77 to 61, with a corresponding reduction in the affected population from 2.35 million (5.5%) to 2.02 million (4.6%).^7^ The number of dynamic municipalities, defined by their economic vitality and medium to high performance in education and longevity indicators, increased from 104 to 112, expanding the share of the population in this category from 32.1% (13.7 million) to 34.0% (15 million). Although the number of unequal municipalities, which generate wealth but exhibit low performance in either education or longevity, rose from 71 to 75, the associated population declined slightly from 45.3% (19.33 million) to 43.6% (19.18 million). Lastly, the group of equitable municipalities, which present low levels of wealth but relatively favourable outcomes in education and longevity, increased from 212 to 218 municipalities, encompassing 9.8% (4.31 million) of the state*’*s population in 2018, compared to 9.7% (4.14 million) in 2014.^7^

### DATA EXTRACTION

The health and socioeconomic information used in this study was obtained from digitised and anonymised data provided by official state agencies. The dependent variable (DV) corresponds to the number of incident cases of leprosy reported in the state of São Paulo during the period from 2017 to 2020. These data were obtained from the Technical Division of Epidemiological Surveillance of Leprosy, part of the Centre for Epidemiological Surveillance of the São Paulo State Health Department.

The independent variables include socioeconomic indicators derived from the São Paulo IPRS, which encompasses municipal-level measures of wealth, longevity, and education. These indicators were obtained from the State Data Analysis System (SEADE Foundation), an agency under the Government of the State of São Paulo. All IPRS data refer to the year 2018 and are publicly available for the state*’*s 645 municipalities.

### COVARIATE SELECTION: MUNICIPAL WEALTH INDICATOR

Among the variables considered in the statistical modelling, the municipal wealth indicator (2018), derived from the IPRS, was selected as a covariate in the Poisson regression model. The choice of this variable was based on both theoretical relevance and empirical significance. The wealth indicator captures the economic profile of each municipality by integrating multiple components. These include electricity consumption in agriculture, commerce, and services; per capita value added; residential electricity consumption; and the average income of formally employed individuals in both the private and public sectors. These data are sourced from administrative records provided by the State Secretariats of Finance and Energy of São Paulo as well as the Ministry of Labour and Employment. The indicator is scaled from 0 to 100 and can be reproduced annually, ensuring its applicability in longitudinal studies.

In the Poisson regression model, Wealth (2018) demonstrated a statistically significant and inverse association with the outcome variable (estimate = –0.03878; *p* < 0.001). This finding suggests that municipalities with higher levels of economic wealth tend to present lower expected counts of incident leprosy cases. This relationship is consistent with prior literature linking socioeconomic development to improved public health and environmental outcomes, reinforcing the theoretical justification for its inclusion in the model. The other covariates, longevity and education, were also evaluated; however, only education showed a statistically significant positive association in the model (estimate = 0.03811; *p* < 0.001), while longevity was not statistically significant (*p* = 0.83). These results inform the understanding of how multidimensional socioeconomic factors influence the phenomenon under study.

### SPATIAL MODELLING: BYM2 MODEL

In this study, we applied the BYM2 model to estimate the spatial distribution of leprosy incidence across municipalities in the state of São Paulo. Although standardised incidence ratios (SIRs) are commonly used in spatial epidemiology, they can be unstable and misleading, particularly in areas with small populations or where the disease is rare. In such cases, the expected number of cases may be very low, leading to high variance and potential misinterpretation of risk patterns. To address this limitation, hierarchical Bayesian models such as the BYM2 offer a more robust alternative by borrowing information from neighbouring regions and integrating relevant covariates. This enables the smoothing of extreme values and enhances the stability of risk estimates.^8,9^

Simpson *et al*.^10^ proposed the BYM2 model as an updated and reparameterised version of the traditional Besag–York–Mollié (BYM) model.^11^ It enables the specification of interpretable and computationally efficient penalised complexity (PC) priors. In this model, the relative risk for area iii, denoted as ‘λi’, is modeled as:

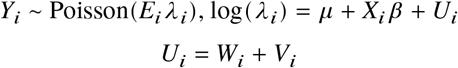

where *E*_*i*_ is the expected number of cases in area *i, X*_*i*_*β* represents the fixed effects of covariates (e.g., municipal wealth), and *U*_*i*_ is a spatially structured random effect combining *W*_*i*_, an intrinsic conditional autoregressive (CAR) term, and *V*_*i*_, an unstructured independent noise term. In the BYM2 formulation, these components are scaled and combined into a single expression to balance structured and unstructured effects via a mixing parameter *ϕ*, as follows:

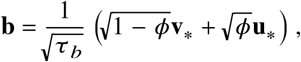

where *τ*_*b*_ is the overall precision (inverse variance) of the combined spatial effect, and *ϕ* ∈ [0,1] controls the proportion of total variance attributed to the structured spatial component **u**_*_. When *ϕ* = 1, the model behaves as a fully structured spatial model, whereas *ϕ* = 0 corresponds to a model driven entirely by unstructured noise.

This reparameterisation simplifies model interpretation and facilitates prior specification in the Bayesian framework. In particular, PC priors can be applied to penalise model complexity, thereby shrinking the structured spatial component toward a base model of spatially unstructured random variation, unless the data strongly support it. The model was implemented using the INLA (Integrated Nested Laplace Approximation) approach in R-INLA. The use of the BYM2 model in this context ensures statistically stable and spatially coherent estimates of leprosy risk, accommodating the heterogeneity in population size and enabling robust inference across small areas.

The marginal precision of the spatially structured random effect (*τ*_*b*_) was assigned a penalized complexity (PC) prior defined as 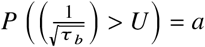. Following the rule of thumb by Simpson *et al*.,^10^ a marginal standard deviation of approximately 0.5 was assumed as a reasonable upper bound and thus set *U* = 0.5/0.31 and *a* = 0.01, yielding the prior 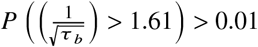. For the mixing parameter *φ*, which balances spatially structured and unstructured effects, we specified the prior *P*(*φ* < 0.5) = 2/3 reflecting a conservative assumption that the unstructured component accounts for more of the variability. Given the presence of 191 zeros in the dataset, we also investigated the potential for zero inflation and selected model configurations accordingly to improve computational efficiency. For transparency and reproducibility, the final model expression below is presented using the syntax implemented in the INLA syntax rather than traditional mathematical notation:

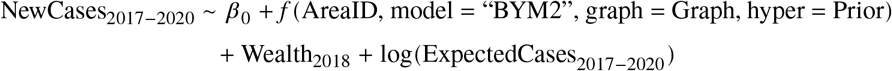

### COMPUTATIONAL STATISTICAL ANALYSIS

Exploratory data analysis was initially performed to detect outliers, missing values and assess zero inflation in the response variable. A spatial neighbourhood map was constructed to identify adjacent municipalities within São Paulo State, defining the neighbourhood structure required for spatial modelling (Figure 1). This adjacency matrix was built using geographic boundary data to determine which municipalities share common borders, establishing the spatial relationships necessary for incorporating spatial dependence in the analysis. One isolated polygon was identified during this process. Spatial neighborhood structures were then defined based on this adjacency information to support spatial modeling, and expected counts were computed for each area *i* using the formula *E*_*i*_ = *r*(*s*) × *n* (*i*), where *r*(*s*) is the overall disease rate (total number of cases divided by the total population across all areas), and *n*(*i*) is the population of area *i*.

**Figure 1.**
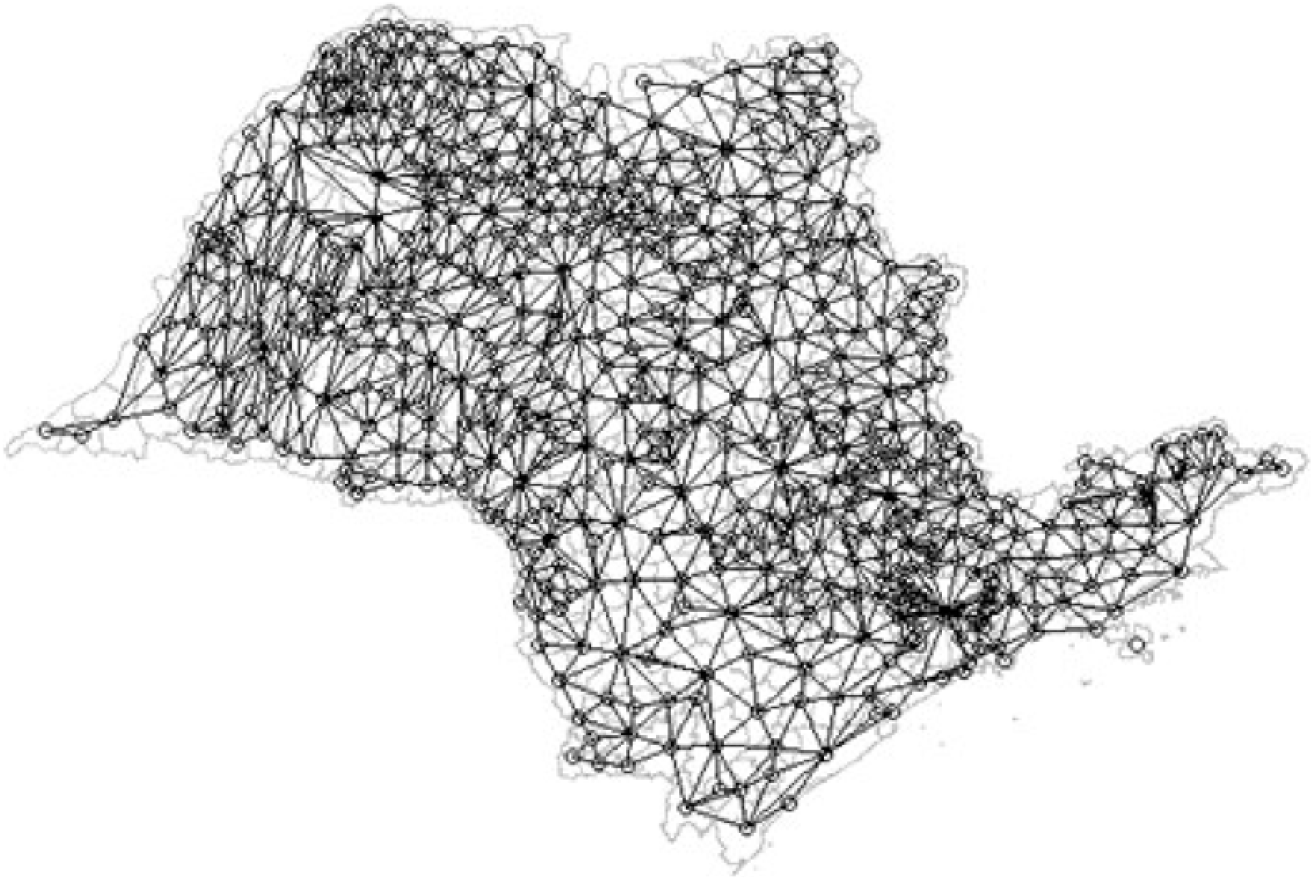
Neighbors of municipalities of São Paulo State.

These expected counts were used to calculate the standardised incidence ratio (SIR) for preliminary spatial risk assessment. Based on exploratory patterns and covariate distributions, final model covariates were selected. The statistical analysis was conducted using R version 4.4.0, with the support of the following packages: *terra, geodata, geostatsp, mapmisc, diseasemapping, INLA*, and *tmap*.

## Results and discussions

To examine the association between socio-territorial indicators and the spatial distribution of leprosy, we analysed municipality-level data for the period 2017–2020 across the 645 municipalities of São Paulo. The analysis combined descriptive epidemiological measures with spatial modelling to identify patterns of risk and their relationship with socioeconomic conditions. During the study period, 4562 new leprosy cases were reported, corresponding to a mean of 7.07 cases per municipality (range: 0–461) and a median of 2 (IQR: 0–5). Expected counts, calculated using the overall state detection rate and municipal population, also averaged 7.07 but ranged widely (0.084–1220.76), reflecting the large variation in population size across municipalities.

The SIR ranged from 0.00 to 32.81 (median 0.82; mean 1.72), indicating that while most municipalities experienced incidence close to or below the expected level, a smaller subset exhibited markedly elevated risk relative to the state average. The municipal wealth index (2018) showed moderate socioeconomic variability across municipalities (mean 34.6; range 15–62).

To facilitate interpretation of spatial patterns, three complementary maps were generated (Figure 2): observed case counts, expected counts, and SIRs. For interpretation, Panel A illustrates the absolute burden of reported cases across municipalities, whereas Panel B adjusts for population size by presenting the number of cases expected if the state detection rate were uniformly applied throughout the study area. Panel C displays the SIR, enabling identification of municipalities where the observed incidence is higher or lower than expected relative to the state average. Geographically, the highest absolute numbers of reported cases in Panel A are concentrated in the more densely populated metropolitan and urbanised areas, particularly in the eastern portion of the state, including the metropolitan region of São Paulo, which largely reflects population concentration. Elevated case counts are also observed in the northern region (Fernandópolis municipality) and in the northwestern region around São José do Rio Preto and Ribeirão Preto, where case counts range from approximately 100 to 461. These municipalities are generally characterised by larger populations, which partly explains the higher number of reported cases, although Fernandópolis represents a notable exception, with relatively elevated case counts despite its smaller population size. Similarly, Panel B shows a spatial pattern driven by demographic distribution, with larger expected counts occurring in municipalities with greater population size across the eastern and central regions of São Paulo. However, Panel C reveals a different pattern when incidence is standardised: several municipalities in the western and northwestern regions exhibit SIR values above the state average, indicating higher-than-expected incidence relative to population size. These areas, therefore, represent atypical patterns in which the disease burden cannot be explained solely by population density and may reflect localised epidemiological or socio-territorial determinants of transmission. This comparison helps distinguish areas with truly elevated risk from those where high case numbers simply reflect larger population size.

**Figure 2.**
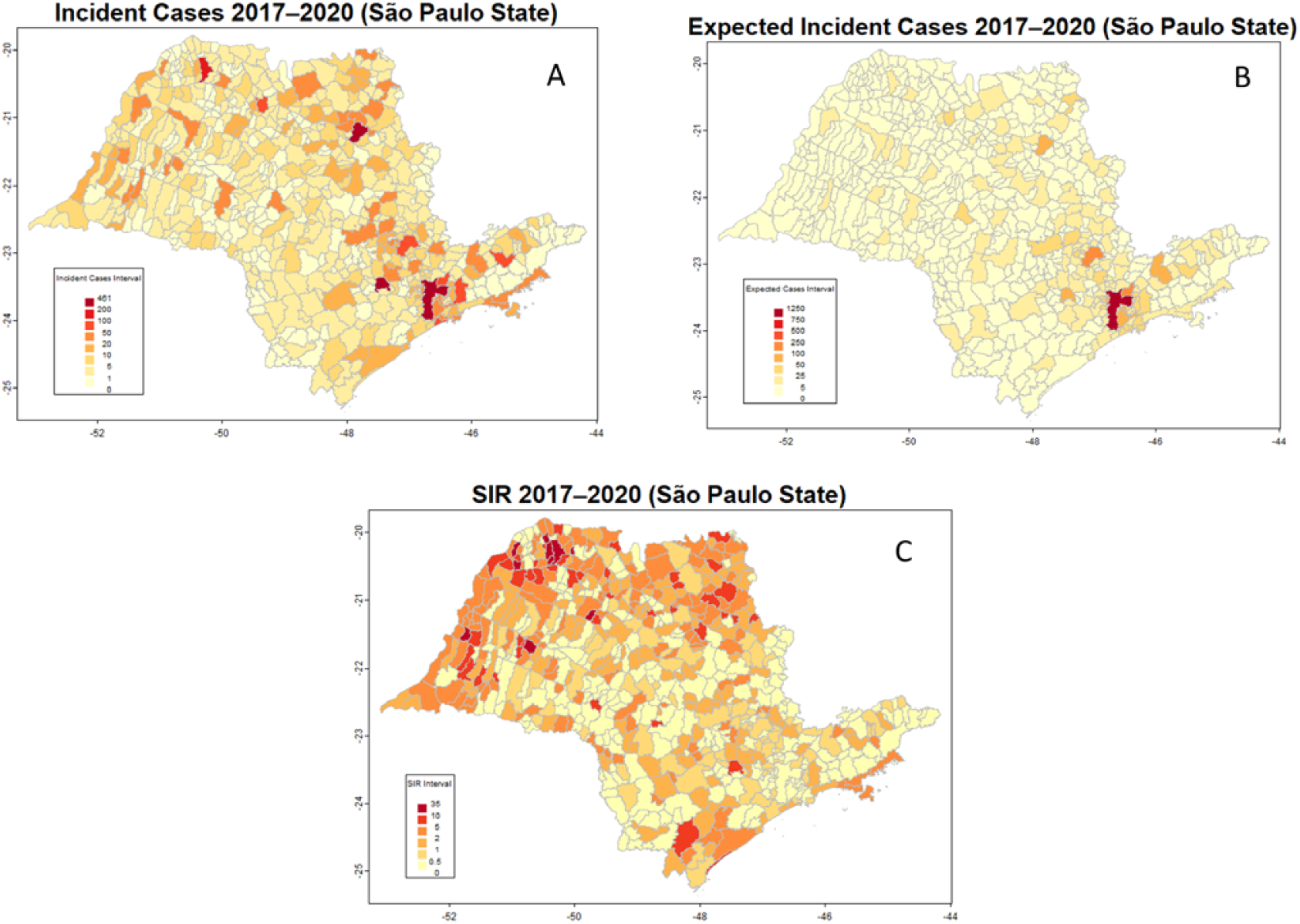
Spatial distribution of observed cases (A), Expected cases (B), and SIRs (C) for Leprosy (2017–2020) in São Paulo State.

While these descriptive maps provide an overview of spatial variation, model-based estimates are necessary to account for spatial dependence and stabilise estimates in municipalities with small populations.

To quantify the association between municipal socioeconomic conditions and leprosy incidence, a spatial Bayesian model using the BYM2 specification was fitted (Table 1).

**Table 1.** Posterior means and 95% credible intervals for model parameters obtained from fitting the São Paulo leprosy data with the BYM2 model Correspondence to: Carlos Souto dos Santos Filho © The author(s). This article is Open Access under CC BY 4.0 1

|  | Mean | 0.025 quantile | 0.975 quantile |
| --- | --- | --- | --- |
| Intercept | 1.138 | 0.362 | 1.915 |
| Wealth | -0.059 | -0.081 | -0.037 |
| Precision ($\tau$) | 0.430 | 0.350 | 0.522 |
| Phi ($\varphi$) | 0.517 | 0.349 | 0.684 |

The estimated intercept (log-risk) was 1.138 (95% CrI: 0.362–1.915). Municipal wealth showed a statistically significant negative association with leprosy incidence (β = *−*0.059; 95% CrI *−*0.081 to *−*0.037), indicating that municipalities with higher socioeconomic status tended to have lower relative risk.

The precision parameter (*τ* = 0.430; 95% CrI 0.350–0.522) and the mixing parameter (*φ* = 0.517; 95% CrI 0.349–0.684) suggest that both structured spatial dependence and local unstructured heterogeneity contribute to the observed variation in risk.

The posterior distribution of the wealth coefficient is illustrated in Figure 3. The entire 95% credible interval lies below zero, providing strong evidence for a negative association between wealth and leprosy risk.

**Figure 3.**
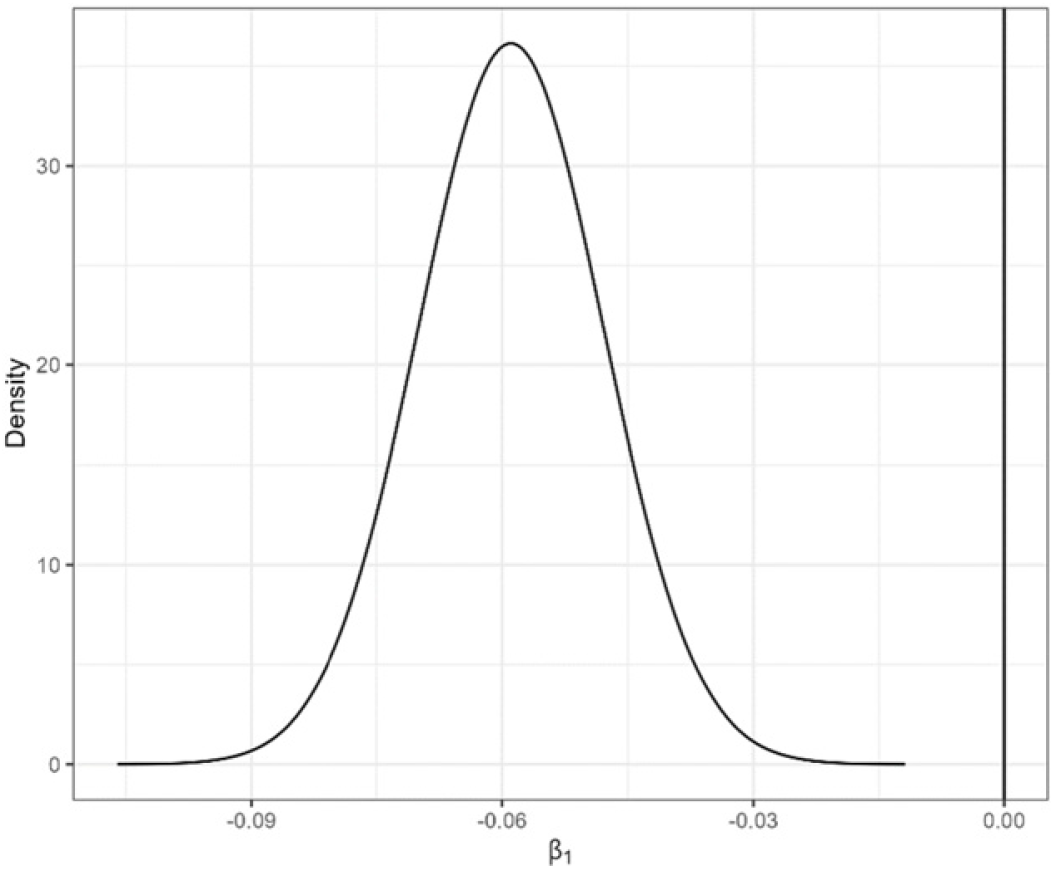
Posterior distribution of the coefficient of covariate wealth.

Posterior municipal relative risks (RR) had a median of 0.96 and a mean of 1.75, with values ranging from 0.06 to 31.52 (Table 2). Figure 4 presents the spatial distribution of the posterior mean RR and its uncertainty. Municipalities with darker shading in Panel A represent areas with higher estimated risk, while Panels B and C show the lower and upper limits of the 95% credible interval, highlighting areas where uncertainty is greater. In the colour scale used in the maps, darker purple tones represent lower RR values, while progressively lighter colours correspond to higher values, allowing areas with elevated estimated risk to be visually distinguished from municipalities with lower risk. The lower limit (LL) of the 95% credible interval varied from 0.0009 to 21.92, while the upper limit (UL) ranged from 0.35 to 53.10. These values indicate that although the majority of municipalities had low or average risk, there were high-risk clusters with substantially elevated relative risk, suggesting spatial heterogeneity in the distribution of leprosy that may be associated with underlying socio-environmental determinants.

**Table 2.** Summary statistics for RR, LL, and UL variables.

| Statistic | RR | LL | UL |
| --- | --- | --- | --- |
| Min. | 0.06095 | 0.000872 | 0.3514 |
| 1st Qu. | 0.46445 | 0.048239 | 1.5544 |
| Median | 0.96117 | 0.217093 | 2.9310 |
| Mean | 1.74752 | 0.648540 | 4.5265 |
| 3rd Qu. | 1.94335 | 0.568785 | 5.8618 |
| Max. | 31.51820 | 21.916249 | 53.0994 |

**Figure 4.**
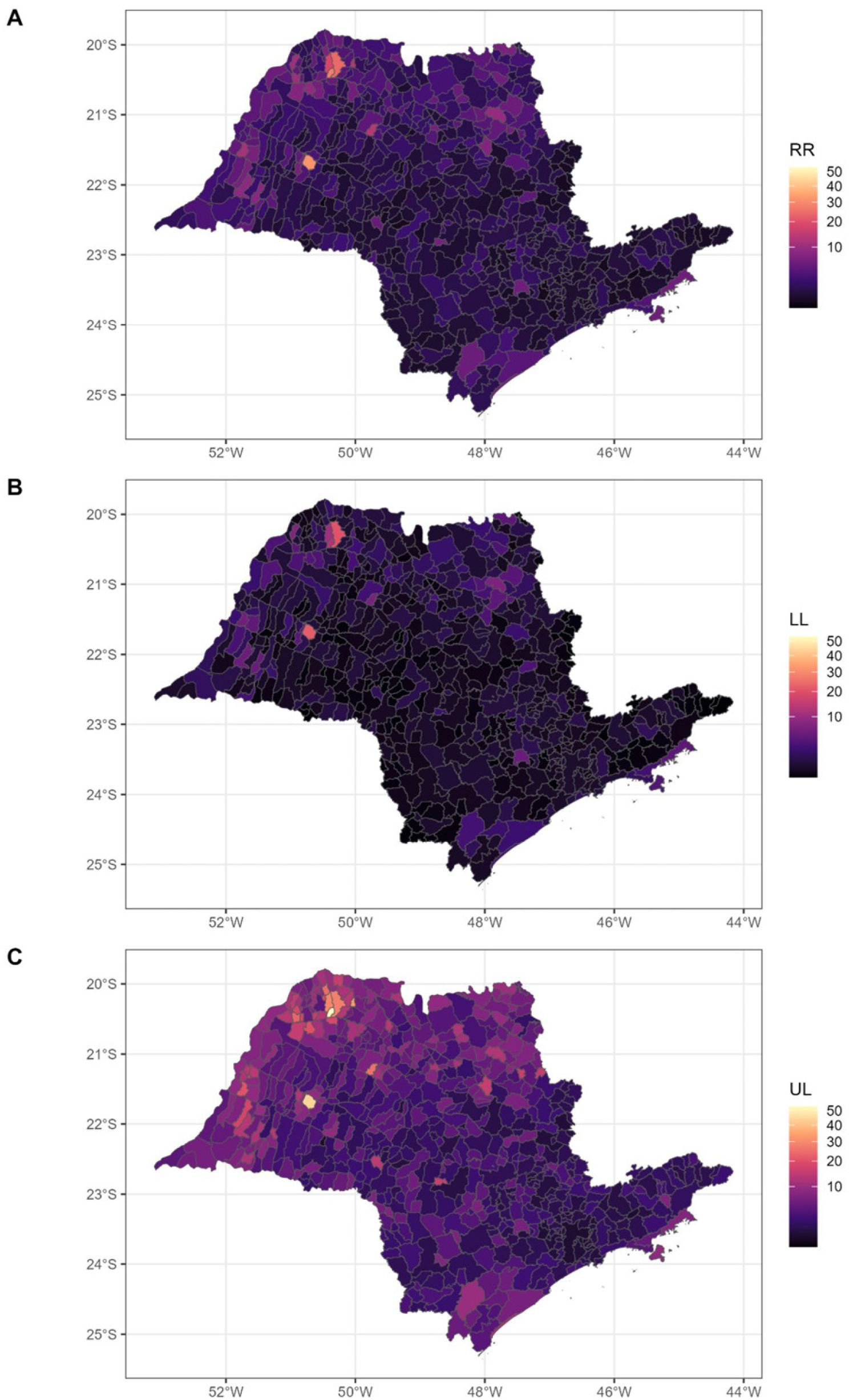
Mean (A) and lower (B) and upper (C) limits of 95% credible intervals of leprosy RR in São Paulo municipalities.

To further understand the spatial heterogeneity of leprosy risk beyond observed covariates, we examined the posterior mean of the spatial random effect *b* from the BYM2 model. This parameter captures unmeasured spatial variation by combining both structured (spatially correlated) and unstructured (random noise) components. Figure 5 presents the posterior mean estimates of *b* across all municipalities in São Paulo. Positive values of *b* indicate areas with higher-than-expected leprosy risk not explained by the included covariate (wealth), while negative values suggest reduced risk or potential underreporting. This spatial residual helps identify hidden patterns of disease transmission and suggests municipalities where additional epidemiological investigation may be warranted. Importantly, this interpretation is strengthened when the spatial random effects are considered together with other model outputs, such as the estimated association with wealth and the predicted municipal relative risks, since the combined set of results provides a more comprehensive picture of spatial risk patterns than observed data or latent effects alone.

**Figure 5.**
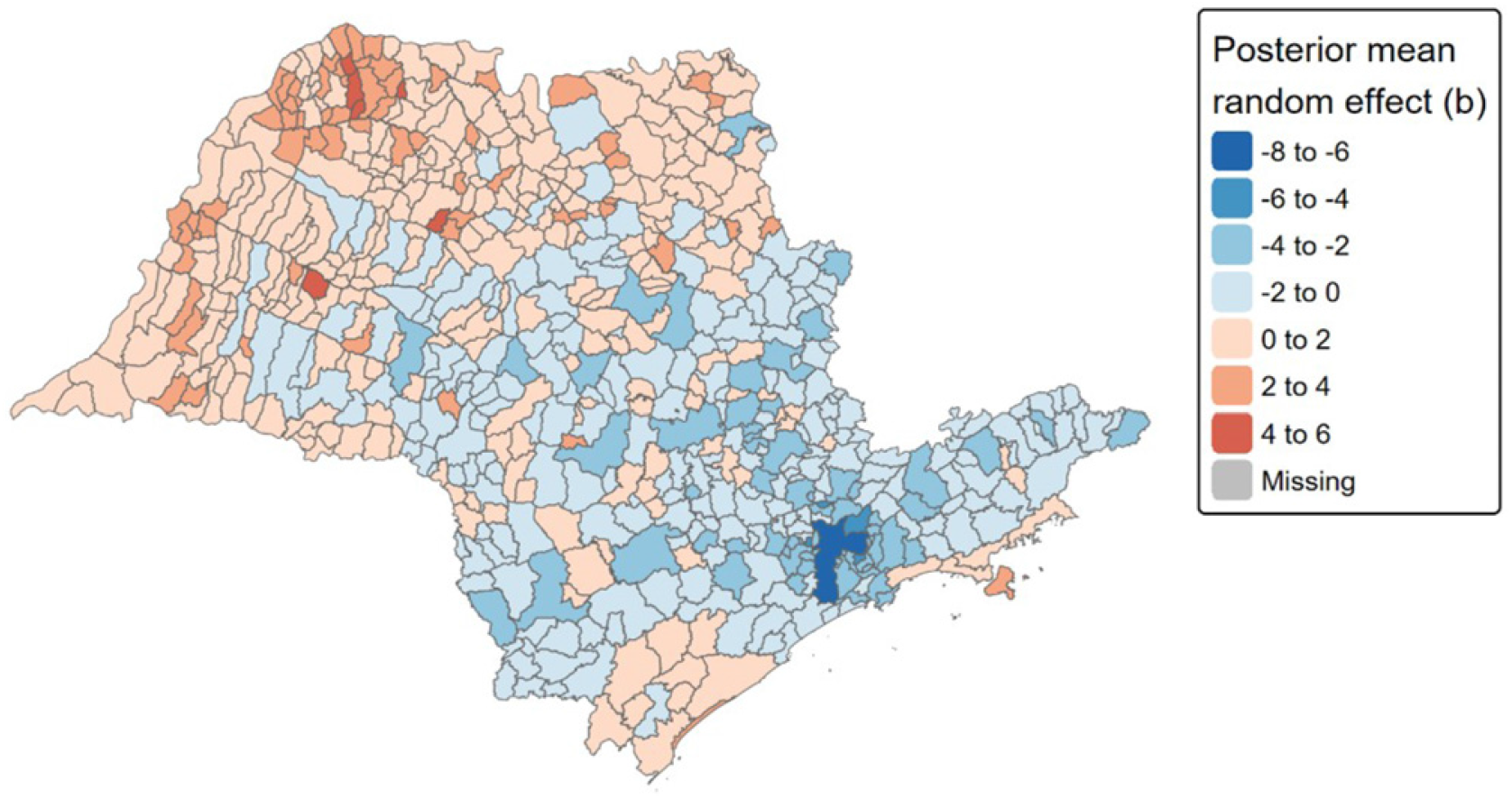
Posterior mean of the BYM2 random effect *b*.

To identify areas with significantly elevated leprosy risk, exceedance probabilities were calculated using a threshold of RR > 2, a criterion commonly adopted in public health to flag high-priority regions. This threshold reflects municipalities where the relative risk is at least double the state average, indicating potential hotspots of concern.

Figure 6 displays the spatial distribution of exceedance probabilities across São Paulo municipalities. The colour scale aids interpretation: light yellow areas represent low probabilities (low-risk), orange to red areas indicate moderate evidence of elevated risk, and dark red to purple areas signal high probabilities (close to 1), reflecting strong statistical evidence that RR exceeds 2.

**Figure 6.**
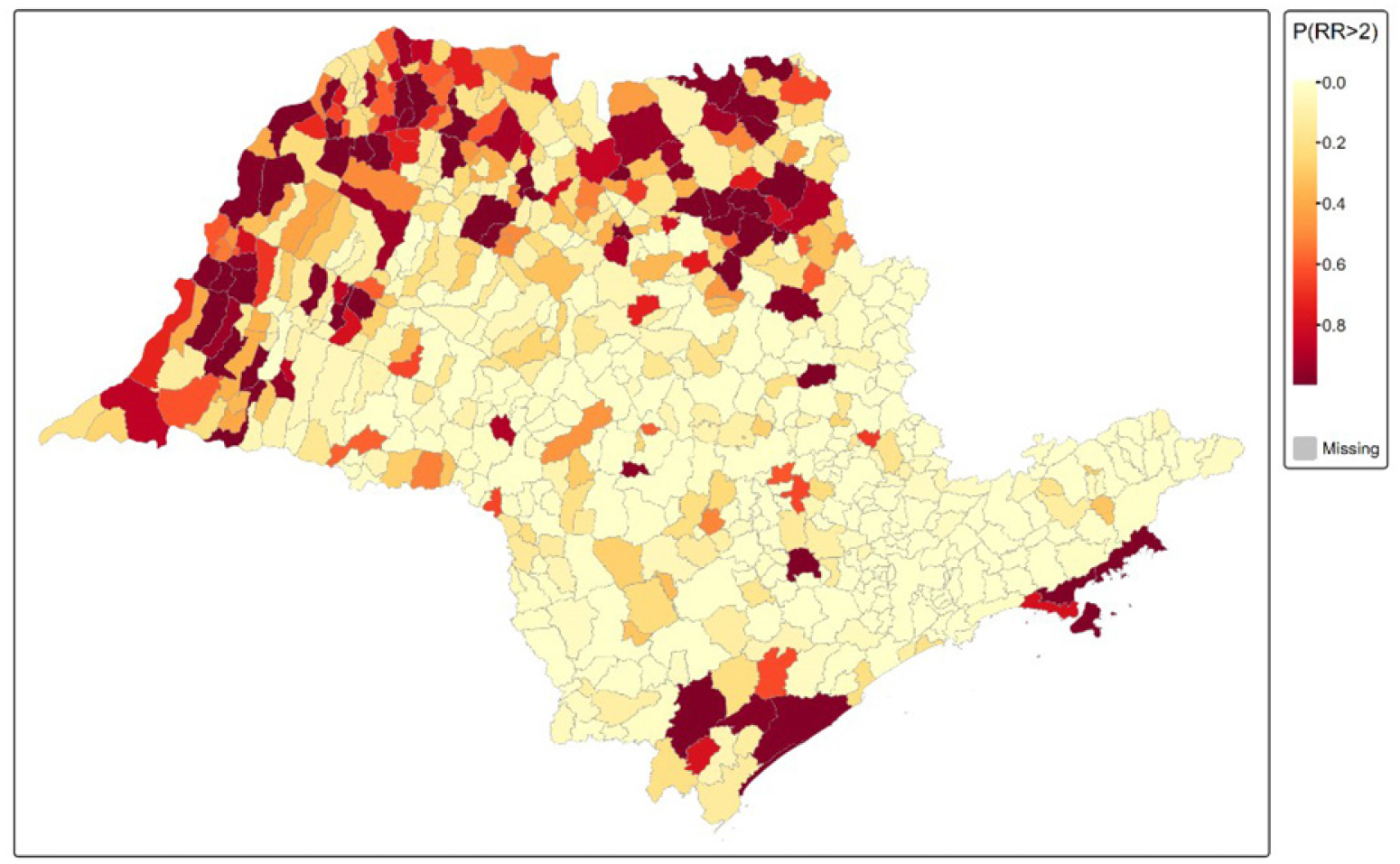
Exceedance probabilities for leprosy relative risk greater than 2 across municipalities in São Paulo (2017–2020).

The map reveals distinct spatial patterns. Clusters of high exceedance probabilities are concentrated in the western and northern regions of the state, while southeastern coastal areas also show some persistent high-risk zones. In contrast, central and southern interior municipalities generally display low probabilities, suggesting a lower likelihood of elevated leprosy risk.

To assess the quality of the fitted BYM2 spatial model, posterior samples and densities were extracted using INLA’s internal sampling functions (*n* = 1000). This approach enables visualisation of marginal posterior distributions and contributes to evaluating model fit beyond summary statistics (Figure 7). The extracted densities help identify the distributional behaviour of key parameters and verify whether the posterior estimates are consistent with prior assumptions and data structure. It is worth noting that the R package used for model fitting and visualisation does not natively support the simultaneous plotting of prior and posterior distributions. Therefore, a direct visual comparison of how the data updates the prior distribution for each parameter could not be incorporated into Figure 7.

**Figure 7.**
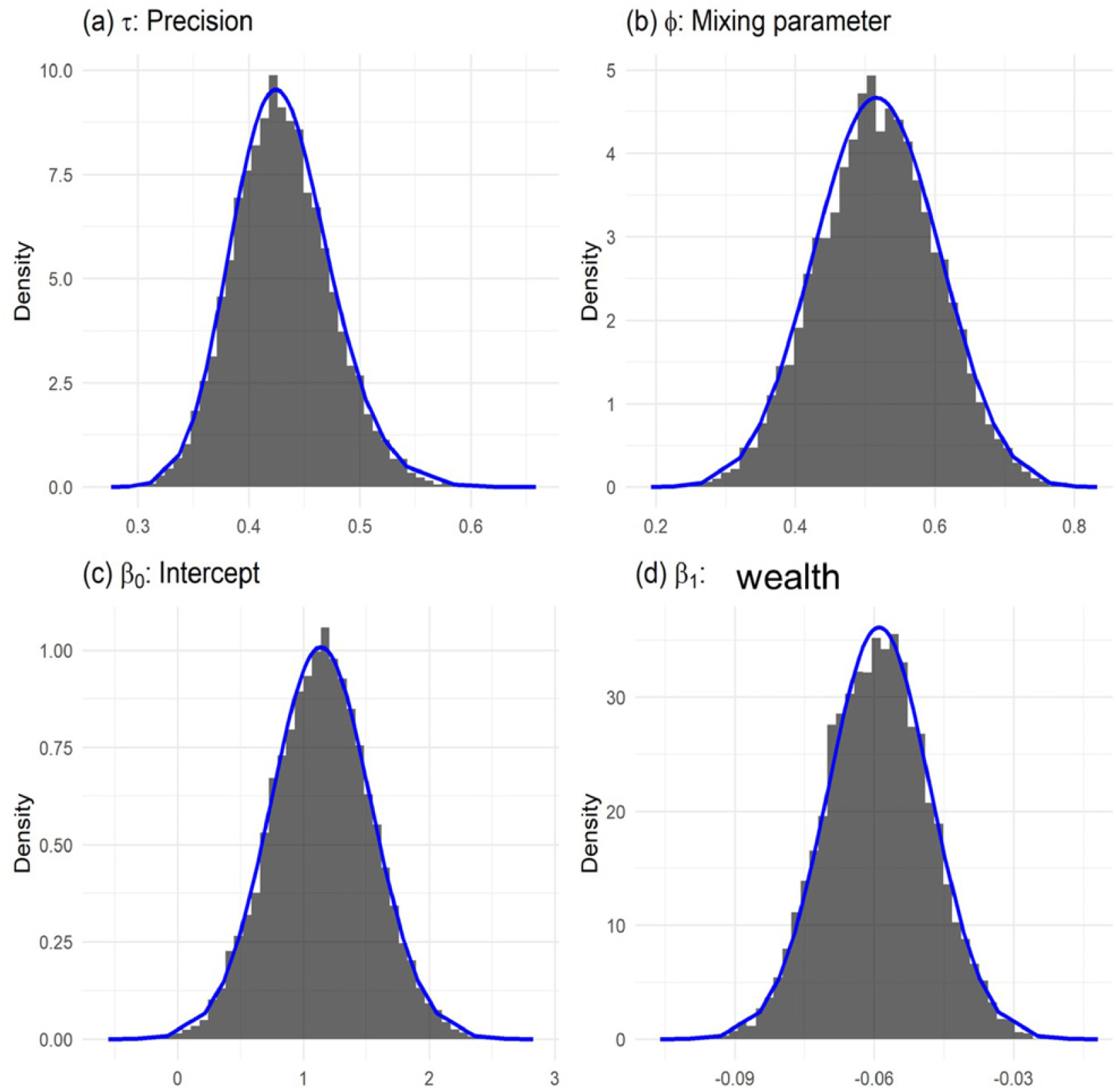
Histograms of posterior samples (black bars) obtained from fitting the BYM2 model using INLA (*n* = 1000), along with posterior densities derived from the marginal distributions (blue lines).

Model fit was further assessed using the Deviance Information Criterion (DIC). The DIC value of 2526.27 reflects the balance between model fit and complexity; although not interpretable in isolation, it serves as a basis for model comparison. The Conditional Predictive Ordinate (CPO) values were also examined to evaluate predictive performance and potential outliers. The distribution of CPO values, where 98% (633 out of 645) of municipalities had CPO < 0.01, suggests low predictive accuracy in many areas. This pattern may indicate overdispersion, zero inflation, or deviation from the assumed Poisson structure in specific municipalities. These findings underscore the need to interpret spatial patterns with caution, particularly in locations with extreme or sparse data.

The BYM2 model revealed significant spatial heterogeneity in leprosy risk across the 645 municipalities, with relative risk estimates ranging from 0.06 to 31.52. Municipal wealth emerged as a significant protective factor (posterior mean: –0.059, 95% CrI: –0.081 to –0.037), indicating that economically advantaged areas consistently demonstrated lower leprosy incidence rates. The mixing parameter (*φ* = 0.517) suggested approximately equal contributions of structured spatial dependence and unstructured random variation in explaining disease distribution. Exceedance probability analysis identified distinct high-risk clusters primarily concentrated in western and northern regions of the state, where municipalities exhibited probabilities exceeding the threshold of RR greater than 2.0, representing areas with at least double the state average incidence rate.

The observed spatial clustering of leprosy cases reflects persistent socioeconomic disparities and suggests the influence of unmeasured environmental or social determinants operating at the municipal level. The concentration of high-risk areas in western and northern São Paulo aligns with historical patterns of regional development inequality, where these areas typically exhibit lower infrastructure investment and reduced access to specialised healthcare services. The stability of these spatial patterns over the four-year study period indicates sustained exposure conditions rather than temporary epidemiological fluctuations, similar to findings in other chronic infectious diseases where poverty-related factors create enduring transmission environments. The negative association between municipal wealth and leprosy incidence supports the well-established relationship between socioeconomic development and infectious disease burden, suggesting that comprehensive poverty reduction strategies may serve as effective primary prevention measures. Additionally, the substantial unexplained spatial variation captured by the random effects component indicates that factors beyond municipal wealth, such as housing quality, population density, healthcare accessibility, or cultural practices, likely contribute to disease transmission dynamics and warrant further epidemiological investigation.

This study demonstrates several methodological strengths, including the use of four years of comprehensive surveillance data from São Paulo’s robust epidemiological system, ensuring high case ascertainment and diagnostic reliability. The BYM2 modelling approach effectively addressed the statistical challenges inherent in rare disease mapping by borrowing strength from neighbouring municipalities and incorporating spatial dependence, thereby producing more stable risk estimates than traditional standardised incidence ratios. The integration of penalised complexity priors prevented overfitting while maintaining interpretability, because these priors constrain the spatial random effects toward a simpler base model (i.e., no spatial structure) unless the data provide sufficient evidence for additional complexity. In the context of our municipal data, where several areas report few cases, this approach reduces the risk of spurious spatial patterns while still allowing meaningful spatial variation to be detected. The exceedance probability framework provided a practical tool for public health priority setting. However, important limitations must be acknowledged. The use of municipal-level aggregated data may mask within-municipality heterogeneity and introduce ecological fallacy when inferring individual-level relationships. Residence at diagnosis served as a proxy for exposure location, potentially misclassifying individuals with complex migration histories or long latency periods typical of leprosy. The model’s limited covariate structure, while statistically significant, explained only a portion of the observed spatial variation, suggesting that crucial unmeasured factors, including individual-level sociodemographic characteristics, environmental exposures, and healthcare utilisation patterns, remain unaccounted for. Furthermore, the high proportion of municipalities with low predictive accuracy (CPO < 0.01 in 98% of areas) indicates potential model misspecification or overdispersion that may affect the reliability of risk estimates in specific locations.

## Conclusions

The observed distribution of leprosy incidence in São Paulo State exhibits spatial clustering associated with municipal wealth disparities. The persistent geographic patterns suggest that socioeconomic conditions are associated with variations in local leprosy risk across municipalities. These findings support targeted public health interventions in identified high-risk areas and highlight the importance of addressing underlying sociodemographic inequalities that may influence the local effectiveness of leprosy control strategies. Future research should explore approaches that integrate high-resolution spatial and temporal data with analyses stratified by vulnerable groups through spatio-temporal modelling frameworks to better understand disease dynamics and inform precision public health interventions.

## Data Availability

All data produced in the present study are available upon reasonable request to the authors

## Ethical considerations

Regarding research ethics and risk assessment, it is important to highlight that the study poses no risk to participants, since all data were previously anonymised and only epidemiologic, non-sensitive information was accessed. Therefore, the requirements of the Brazilian General Data Protection Law (LGPD - Law No. 13.853/2019) were fully met, which specifically provides for the possibility of processing data for scientific purposes without the need for the data subject’s consent.

## Authors’ contributions

CSSF conceived the study; CSSF designed the study protocol; CSSF collected data, performed data entry and carried out the data analysis and interpretation; CSSF, AJAC and GASA drafted the manuscript; CSSF, AJAC and GASA critically revised the manuscript for intellectual content; CSSF, AJAC and GASA read and approved the final manuscript. CSSF is guarantor of the paper.

## Patient consent statement

No patient consent was required.

## Data sharing statement

The supporting data for this article can be obtained from the corresponding author on request.

